# Genetic crossroads of cardiovascular disease and its comorbidities: Toward holistic therapeutic strategies

**DOI:** 10.1101/2025.06.17.25329808

**Authors:** Binisha H. Mishra, Pashupati P. Mishra

## Abstract

With increasing life expectancy, the prevalence of cardiovascular disease (CVD) accompanied by comorbidities is rising, presenting a growing challenge for healthcare systems. Understanding shared genetic factors underlying CVD and its comorbidities can help develop more effective prevention and treatment strategies. In this study, we investigated genetic correlations between CVD and common comorbidities using genome-wide association study (GWAS) summary statistics from the FinnGen R12 release. Following standard quality control procedures, we examined 19 disease endpoints using linkage disequilibrium score regression (LDSC) to estimate heritability and pairwise genetic correlations. Disease traits with significant heritability (z-score ≥ 4) and Bonferroni-corrected significant correlations (adjusted p < 0.05) were selected for genomic structural equation modeling (Genomic SEM) to construct a latent genomic factor (LGF), representing shared genetic liability. Out of the 19 diseases, four CVDs (transient ischemic attack, atrial fibrillation, myocardial infarction and heart failure) and seven comorbidities (type 2 diabetes, asthma, obesity, depression, chronic obstructive pulmonary disease, gingivitis and hypertension) showed statistically significant genetic correlations. A multivariate GWAS of the LGF identified 141 novel associated loci across 29 independent SNPs. These loci overlapped with 16 protein-coding genes, including *NPC1, TMEM106B, PTPN22, MAP2K5 and MSRA*, implicating them in the shared pathogenesis of CVD and its comorbidities. These findings highlight a shared genetic architecture underlying CVD and its comorbidities, revealing cross-disease genetic risk factors that may enhance joint risk prediction, inform precision medicine strategies, and offer insights into common biological mechanisms and potential targets for integrated therapeutic approaches.

## 1. Background

Cardiovascular diseases (CVDs) remain one of the most urgent global public health challenges, with significant implications for both human well-being and economic stability. In the European Union (EU), CVDs are the leading cause of mortality, accounting for approximately 32.4 % of all deaths [Eurostat, 2023]. The economic burden is equally alarming, with an estimated annual cost of around €282 billion, driven by healthcare expenses, lost productivity, and informal care [Luengo-Fernandez et al., 2023]. Despite global initiatives aimed at reducing their impact, progress in mitigating the burden of CVDs has been inadequate. At the same time, the prevalence of comorbid chronic conditions alongside CVDs continues to rise. This trend has led the European Commission to recognize comorbidity as a key priority, promoting awareness and fostering innovation in healthcare solutions to better address the growing complexities of managing multiple chronic conditions [European Commission, 2015].

There is a growing consensus among public health experts and policymakers that healthcare systems must shift away from a narrow, single-disease focus toward a more holistic, multimorbidity-oriented approach. Effective management of complex cases involving CVD and its comorbidities requires a deeper understanding of how chronic diseases interact, how they jointly influence health outcomes, and how they affect treatment effectiveness and healthcare utilization. Managing CVD in isolation often neglects the roles of co-occurring conditions such as diabetes, obesity, and depression, which can complicate care and reduce treatment efficacy.

It is crucial to move beyond clinical observations and investigate the underlying biological mechanisms that drive the co-occurrence of these conditions to develop more effective and targeted strategies. While previous studies have explored comorbid relationships between atherosclerosis and individual conditions such as osteoporosis [Mishra et al., 2022] or depression [Mishra et al., 2024], comprehensive investigations that simultaneously consider multiple CVDs and their common comorbidities remain rare. One promising avenue for advancing this understanding is through the exploration of shared genetic architecture, which can reveal underlying biological connections between seemingly distinct CVD and its comorbidities.

A recent study by [Zhao et al., 2024] examined a similar multimorbidity concept using a genome-wide association study (GWAS) approach on cardiometabolic multimorbidity, defined as the coexistence of two or three cardiometabolic diseases. However, the study was based on traditional GWAS methods and did not analyze shared genetic architecture of the CVDs and their comorbidities using multivariate approach. In contrast, the present study aims to assess the genetic correlations between CVD and its most prevalent comorbidities, identify shared genetic factors contributing to their co-occurrence and identify genetic variants associated with this common underlying genetic foundation utilizing advanced statistical methodologies. A deeper understanding of these shared genetic underpinnings of CVD and its comorbidities may uncover common biological pathways and guide the development of more integrated and effective strategies for prevention, treatment, and management.

## 2. Methods

This study was based on GWAS summary statistics of 19 clinical endpoints related to CVDs and their comorbidities from FinnGen data freeze 12 [Kurki et al., 2023]. Clinical endpoints were constructed from the registry codes utilizing the Finnish adaptation of the International Classification of Diseases, 10th revision (ICD-10) diagnosis codes, while aligning them with the definitions provided in ICD-8 and ICD-9 [Supplementary Table 1]. The FinnGen study is a large-scale genomics initiative that has analyzed over 500,000 Finnish biobank samples and correlated genetic variation with health data to understand disease mechanisms and predispositions. The project is a collaboration between research organizations and biobanks within Finland and international industry partners. The number of cases and controls for each disease, along with the effective sample size after correcting case-control imbalance, are presented in Table 1. Selection of GWASs on CVDs and their comorbidities was based on known comorbidities of CVD [Buddeke et al., 2019] that were available in FinnGen database and had sample overlap ranging from 7–76% [Supplementary Table 2].

**Table 1.** Cardiovascular diseases and their associated comorbidities included in this study, with number of cases, controls, effective sample size and genome-wide significant loci identified by univariate genome-wide association studies (GWASs).

|  | **Diseases** | **Cases** | **Controls** | **Effective sample size** | **Significant loci** |
| --- | --- | --- | --- | --- | --- |
| **Cardiovascular comorbidities** |  |  |  |  |  |
| 1. | Coronary atherosclerosis | 63307 | 416171 | 219793.5 | 154 |
| 2. | Transient ischemic attack | 24948 | 453276 | 94586.05 | 2 |
| 3. | Stroke | 34110 | 450023 | 126827 | 10 |
| 4. | Atrial fibrillation | 63532 | 252810 | 203090.6 | 177 |
| 5. | Myocardial infarction | 31666 | 416171 | 117707.7 | 84 |
| 6. | Heart failure | 37653 | 462695 | 139277.9 | 18 |
| 7. | Cardiomyopathy | 7649 | 375343 | 29984.95 | 47 |
| **Non-cardiovascular comorbidities** |  |  |  |  |  |
| 8. | Type 2 diabetes | 55418 | 405261 | 195005.7 | 272 |
| 9. | Hypertension | 154630 | 345634 | 427337.4 | 424 |
| 10. | Asthma | 47300 | 259839 | 160062.8 | 87 |
| 11. | Metabolic dysfunction associated fatty liver disease | 3504 | 496844 | 13917.84 | 7 |
| 12. | Osteoporosis | 10461 | 473264 | 40939.08 | 13 |
| 13. | Obesity | 31499 | 468693 | 118061.6 | 76 |
| 14. | Depression | 59333 | 434831 | 208836.2 | 45 |
| 15. | Chronic obstructive pulmonary disease | 24138 | 409070 | 91172.2 | 35 |
| 16. | Epilepsy | 15645 | 374605 | 60071.19 | 4 |
| 17. | Pulmonary embolism | 12762 | 486319 | 49742.65 | 33 |
| 18. | Rheumatoid arthritis | 16314 | 315115 | 62043.89 | 45 |
| 19. | Gingivitis | 137839 | 310260 | 381754.3 | 2 |

Quality control steps in the GWAS involved exclusion of participants with ambiguous gender, high genotype missingness (>5%), excess heterozygosity (+-4SD) and non-Finnish ancestry. SNPs with high missingness (>2%), low HWE p-value (<1e-6) and low minor allele count (MAC<3) were excluded.

Heritability for each disease and the genetic correlations between them were assessed using linkage disequilibrium score (LDSC) regression [Bulik-Sullivan et al., 2015]. This study included only diseases with a heritability z-score ≥ 4, given that genetic correlation estimates below this threshold are typically characterized by high noise and limited reliability. Genetic correlations with Bonferroni-adjusted p-values (adj.pval) < 0.05 were considered statistically significant.

A latent genetic factor (LGF) was constructed using genomic structural equation modeling (genomic SEM), based on the set of genetically correlated traits by leveraging the genetic covariance structure between them [Grotzinger et al., 2019]. A multivariate GWAS of LGF was conducted using Genomic SEM to find shared genetic architecture of cardiovascular diseases and their comorbidities. Independent loci were identified using PLINK 2.0 clumping procedure (– clump-r2 0.6 –clump-kb 1000) [Chang et al., 2015]. The independent loci were annotated using SNPnexus, a web-based tool for genetic variant annotation using data from public repositories [Oscanoa et al., 2020].

## 3. Results

### 3.1 Genetic correlation between CVDs and their comorbidities

Out of 19 clinical endpoints related to CVD and their comorbidities, we considered associations with an adjusted p-value < 0.05 as statistically significant [Figure 1A, Supplementary Table 3]. In addition, we reported selected endpoints that did not meet this threshold, due to their broader relevance across multiple conditions. Specifically, although gingivitis was not significantly associated with heart failure and myocardial infarction, it showed significant associations with nine other conditions. Similarly, chronic obstructive pulmonary disease was not associated with transient ischemic attack but was significantly associated with all remaining endpoints. Depression was not associated with heart failure but was significantly associated with the other conditions. These findings suggest that, despite the absence of significance for a few endpoints, these diseases remain important due to their consistent associations across most of the clinical outcomes. Consequently, we selected 11 of the 19 diseases for further analysis of shared genetic etiology. These included four CVDs (heart failure, myocardial infarction, arterial fibrillation and transient ischemic attack) and seven comorbidities (type 2 diabetes, gingivitis, chronic obstructive pulmonary disease, depression, obesity, asthma and hypertension). Collectively, the results support a shared genetic etiology underlying clinically distinct conditions.

**Figure 1:**
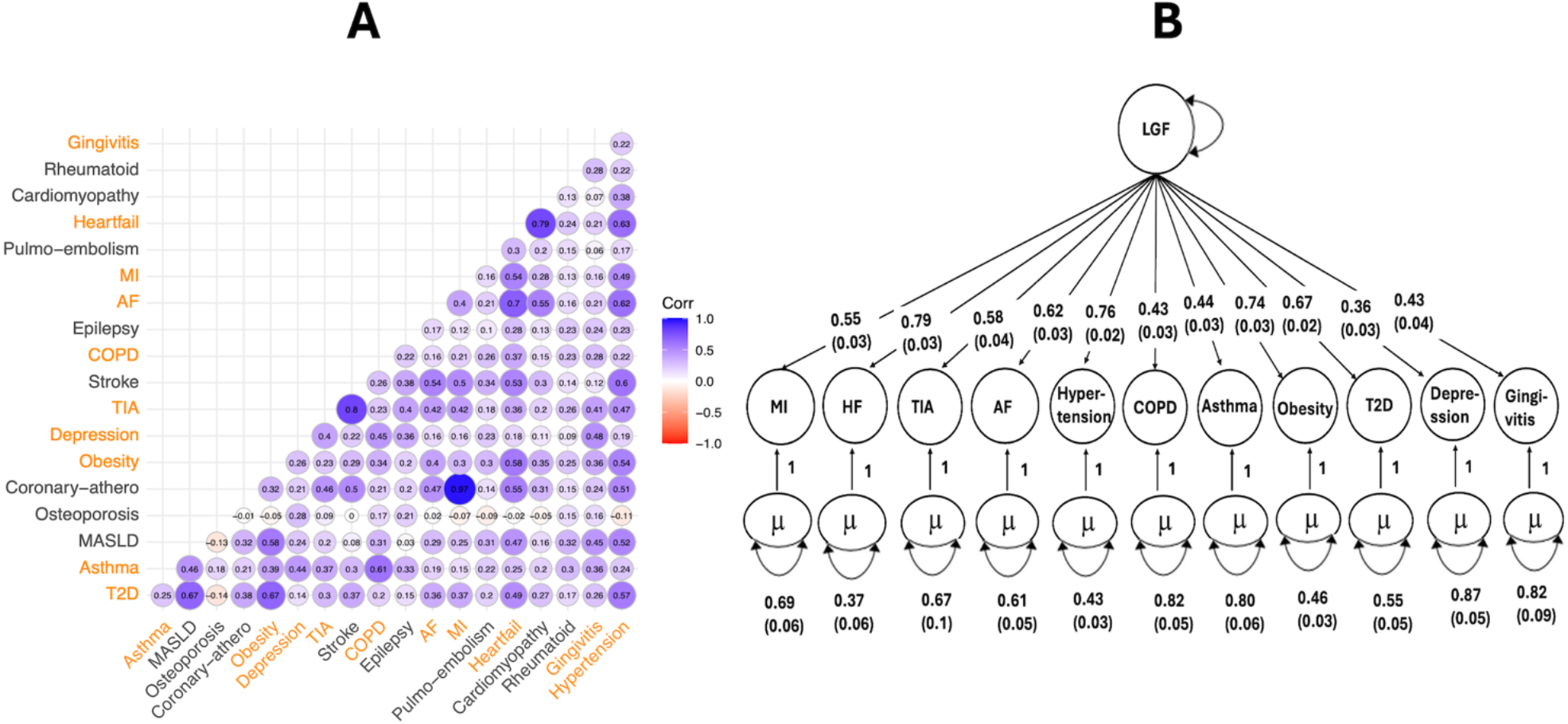
Genetic correlations and multivariate genetic factor model of cardiometabolic diseases. **A**. SNP-based pairwise genetic correlations for the nineteen cardiometabolic disorders estimated using Linkage Disequilibrium Score (LDSC) regression. **B**. Path diagram with standardized factor loadings in the hierarchical model estimated using Genomic SEM. The μ represents the residual variance that is not explained by the latent factor. ***Abbreviations***: Pulmo-embolism, pulmonary embolism; MI, myocardial infarction; AF, atrial fibrillation and flutter; COPD, chronic obstructive pulmonary disease; TIA, transient ischemic attack; Coronary-athero, coronary atherosclerosis; MASLD, metabolic dysfunction-associated steatotic liver disease; T2D, type 2 diabetes mellitus; LGF, latent genetic factor.

**Figure 2:**
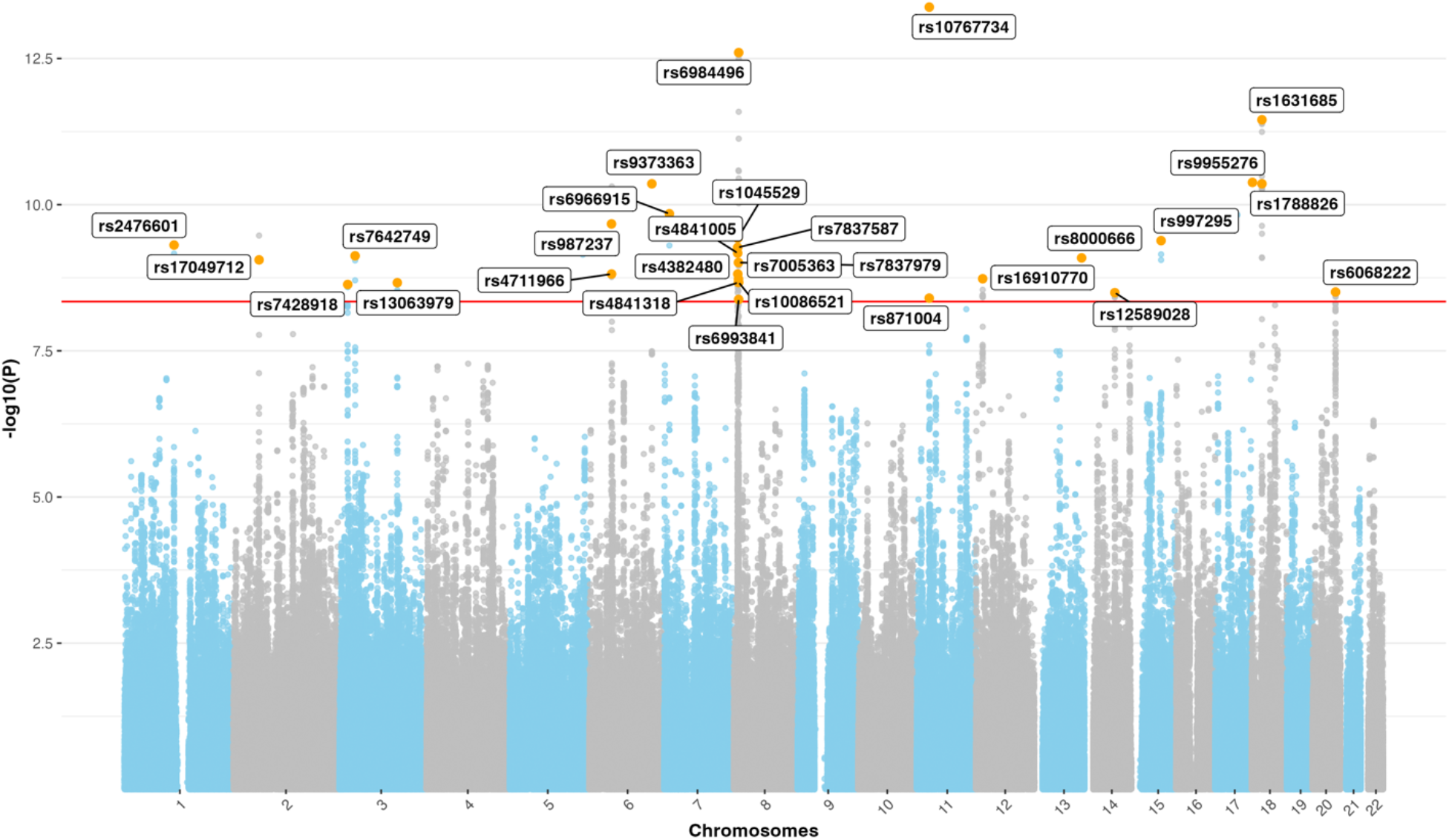
Multivariate genome-wide association study of latent genetic factor (LGF) for cardiovascular diseases and their comorbidities. The x axis represents the chromosomal position, and the y axis represents the −log10 of the unadjusted p-value for SNP associations with linkage disequilibrium score (LDSC) regression.

### 3.2. Shared genetic architecture of CVDs and their comorbidities

Using a multivariate GWAS of the latent genetic factor underlying 11 genetically correlated CVDs and the comorbid conditions, we identified 141 new genome-wide significant SNPs that were not detected in the original univariate GWASs [Table 2]. Of the 141 SNPs, 29 were identified as independent loci through PLINK-based clumping. These 29 SNPs mapped to 16 protein-coding genes involved in several key biological processes associated with CVDs and their comorbidities.

**Table 2.** List of 29 independent novel genetic variants, their corresponding genomic location, p-values and annotation, identified with multivariate genome-wide association study (GWAS) of latent genetic factor (LGF) representing genetically correlated cardiovascular diseases (CVDs) including comorbidities.

| *rsID* | *Chromosome* | *Position* | *P-value* | *Gene* | *Type* |
| --- | --- | --- | --- | --- | --- |
| rs10767734 | 11 | 28620834 | 4.2e-14 | None | None |
| rs6984496 | 8 | 10938583 | 2.5e-13 | XKR6 | protein_coding |
| rs1631685 | 18 | 23554275 | 3.5e-12 | NPC1 | protein_coding |
| rs9955276 | 18 | 1839338 | 4.2e-11 | None | None |
| rs1788826 | 18 | 23574060 | 4.4e-11 | NPC1 | protein_coding |
| rs9373363 | 6 | 142828906 | 4.4e-11 | HIVEP2 | protein_coding |
| rs6966915 | 7 | 12226362 | 1.4e-10 | TMEM106B | protein_coding |
| rs987237 | 6 | 50835337 | 2.1e-10 | TFAP2B | protein_coding |
| rs1045529 | 8 | 9032588 | 3.7e-10 | ERI1 | protein_coding |
| rs997295 | 15 | 67724005 | 4.1e-10 | MAP2K5 | protein_coding |
| rs2476601 | 1 | 113834946 | 4.9e-10 | PTPN22 | protein_coding |
| rs7837587 | 8 | 8521482 | 5.4e-10 | None | None |
| rs4841005 | 8 | 8643720 | 6.7e-10 | None | None |
| rs7642749 | 3 | 35627397 | 7.5e-10 | None | None |
| rs8000666 | 13 | 107712256 | 8.1e-10 | FAM155A | protein_coding |
| rs17049712 | 2 | 58734001 | 8.8e-10 | LINC01122 | lincRNA |
| rs7837979 | 8 | 10341024 | 9.8e-10 | MSRA | protein_coding |
| rs4382480 | 8 | 8863963 | 1.5e-09 | MFHAS1 | protein_coding |
| rs4711966 | 6 | 50996064 | 1.5e-09 | None | None |
| rs16910770 | 12 | 15352797 | 1.8e-09 | PTPRO | protein_coding |
| rs7005363 | 8 | 10426238 | 1.9e-09 | MSRA | protein_coding |
| rs10086521 | 8 | 10926259 | 2.1e-09 | XKR6 | protein_coding |
| rs4841318 | 8 | 10329690 | 2.1e-09 | MSRA | protein_coding |
| rs13063979 | 3 | 131845897 | 2.2e-09 | CPNE4 | protein_coding |
| rs7428918 | 3 | 18782806 | 2.3e-09 | SATB1-AS1 | processed_transcript |
| rs6068222 | 20 | 52541802 | 3.1e-09 | LINC01524 | lincRNA |
| rs12589028 | 14 | 69076025 | 3.2e-09 | DCAF5 | protein_coding |
| rs871004 | 11 | 28490911 | 4.0e-09 | METTL15 | protein_coding |
| rs6993841 | 8 | 10775174 | 4.2e-09 | PINX1 | protein_coding |

These included fundamental cellular functions such as metal ion binding, methyltransferase activity, and protein metabolism; key metabolic processes like the regulation of fatty acid biosynthesis and LDL clearance; as well as pathways involved in calcium ion import across the plasma membrane, signal transduction, immune system function, dendrite morphogenesis, lysosomal trafficking and apoptosis.

## 4. Discussion

In this study, we utilized multivariate genomic approaches to identify novel genetic loci associated with a latent genetic factor underlying 11 genetically correlated cardiovascular diseases (CVDs) and related comorbidities, using GWAS summary statistics from the FinnGen database. The 11 conditions were selected from an initial set of 19 based on significant genetic correlations, indicating a shared genetic architecture. These correlations suggest that the co-occurrence of CVDs and their comorbidities may stem from overlapping genetic mechanisms, beyond shared environmental or behavioral risk factors. With multivariate GWAS of the latent genetic factor, our analysis uncovered genetic loci not previously implicated in individual CVDs, highlighting the value of modeling shared genetic liability. These findings point to common pleiotropic mechanisms contributing to both cardiovascular pathology and its related comorbidities, offering promising avenues for future genetic, biological, and therapeutic investigations.

Functional annotation revealed that many of the identified genes are involved in biological processes relevant to cardiovascular and metabolic health, including lipid metabolism, vascular integrity, mitochondrial function, inflammation, and oxidative stress.

Among the most compelling findings is the association of a SNP within *NPC1* (Niemann-Pick disease, type C1), a gene essential for intracellular cholesterol transport and lipid homeostasis [Trinh et al., 2018]. Dysregulation of NPC1 has been implicated in atherosclerosis [Yu et al., 2014], and its association with the latent genetic factor in our study reinforces its role in lipid trafficking as a shared pathogenic mechanism across genetically correlated CVDs. Similarly, *MSRA* (methionine sulfoxide reductase A), known for its role in reducing oxidative stress via methionine sulfoxide reduction [Moskovitz et al., 2001], has been associated with improved lipid metabolism and atherosclerotic protection in animal models [Xu et al., 2015]. Its known associations with mental health phenotypes [Ni et al., 2015] and personality traits [Boutwell et al., 2017] further suggest a pleiotropic role across cardiometabolic and neuropsychiatric disorders.

The identification of *TMEM106B* (Transmembrane Protein 106B) further underscores the interplay between cardiovascular and neuropsychiatric conditions. Previously linked to both depression and coronary artery disease [Li et al., 2015; Li et al., 2025], *TMEM106B* may represent a therapeutic nexus for comorbid mental and cardiovascular health. A similar pleiotropic pattern emerges with *PINX1* (PIN2 interacting protein 1), implicated in obesity-induced cardiac injury and heart development through its influence on angiogenesis and stem cell differentiation [Pan et al., 2022; Chan et al., 2020].

Our analysis also revealed *PTPN22* (Protein Tyrosine Phosphatase Non-Receptor Type 22), a gene well-known in autoimmune and inflammatory pathways, as a potential contributor to CVD.

Emerging evidence suggests that *PTPN22* plays a role in calcific aortic valve disease (CAVD) pathogenesis, potentially through modulation of immune activity, and its targeted inhibition represents a promising strategy for the development of novel therapeutics against CAVD [Li et al., 2023]. Our findings extend its relevance to broader cardiovascular and inflammatory disease risk, making it a promising therapeutic target.

Genes involved in apoptosis and RNA metabolism also emerged as significant contributors. *XKR6* (XK-related protein 6), predicted to influence apoptotic pathways, is of interest due to apoptosis’s critical role in atherosclerosis progression [Li et al., 2022]. *ERI1* (exoribonuclease 1), associated with carotid intima media thickness [Meena et al., 2025], points to RNA metabolism as a crucial mechanism influencing vascular remodeling and CVD susceptibility.

Inflammation emerged as a common biological link, with *MFHAS1* (Malignant Fibrous Histiocytoma Amplified Sequence 1) and *MAP2K5* (Mitogen-Activated Protein Kinase 5) being key players. *MFHAS1* is involved in regulating inflammation [Ng et al., 2011], and although its link to CVD is not yet well-defined, its roles in neuroinflammation and cognitive decline [Zhong et al., 2019] suggest broader implications for systemic inflammatory processes that intersect with cardiometabolic pathways. Likewise, *MAP2K5*, a component of the *MAPK/ERK5* signaling pathway, influences endothelial dysfunction and atherosclerosis [Johnson & Lapadat 2002; Le et al., 2013]. Targeting *MAP2K5* may offer protective effects against both vascular injury and associated metabolic dysregulation.

We also identified genes that hint at broader systemic interactions. *PTPRO* (protein tyrosine phosphatase receptor type O), which regulates glomerular filtration and blood pressure [Wharram et al., 2000], exemplifies the renal-cardiovascular axis central to hypertension and CVD. *METTL15* (Methyltransferase Like 15), involved in mitochondrial RNA methylation, has been linked to various heart diseases [Kumari et al., 2022], further highlighting the importance of mitochondrial integrity in cardiovascular pathology.

Although some identified genes, such as *HIVEP2* (human immunodeficiency virus type I enhancer binding protein) and *CPNE4* (copine 4), lack direct associations with CVD, their known roles in neurodevelopmental disorders and metabolic regulation [Steinfeld et al., 2016; Goel et al., 2021] suggest potential indirect contributions to cardiometabolic risk, particularly in the context of shared developmental and signaling pathways.

There were certain limitations in our study. First, the effective sample size for each clinical endpoint varied, ranging from 13917,84 for MAFLD to 427337,4 for hypertension and the GWASs of the selected clinical endpoints available in FinnGen database had sample overlap ranging from 7–76%. The varying sample size and overlaps may have impacted our study findings. Also, sex-stratified analysis was not possible due to the use of GWAS summary data as input. Therefore, further research should investigate how shared genetic risks differ by sex. Lastly, all samples were primarily of European ancestry; therefore, our findings are relevant only for European-ancestry populations.

Traditional GWAS approaches often consider diseases in isolation, potentially overlooking the shared biological pathways that drive comorbidity. By modeling a latent genetic factor, we highlight pleiotropic loci that influence multiple related conditions simultaneously. Our findings illustrate a shared genetic framework connecting CVD with diverse comorbid conditions, underpinned by genes involved in lipid metabolism, mitochondrial function, inflammation, apoptosis, RNA regulation, and neurovascular signaling. Several of these genes have previously been proposed as therapeutic targets for individual conditions. Therefore, in addition to deepening our understanding of cardiometabolic and systemic disease etiology, our findings point to a set of potential therapeutic targets such as *NPC1, TMEM106B, PTPN22, MAP2K5*, and *MSRA*, with pleiotropic effects across cardiovascular, metabolic, and neuropsychiatric domains. We provide genetic evidence that these genes should instead be evaluated for their potential in developing holistic, multi-disease therapeutic strategies. Future studies should aim to experimentally validate these findings and explore their translational potential in precision medicine.

## Data Availability

Summary statistics from the FinnGen study are made openly available to the research community following a short embargo period. Access to individual-level data is restricted to research partners and provided within a secure analysis environment, ensuring compliance with data protection regulations and safeguarding participant privacy. Further information on data access procedures is available through the FinnGen website.

https://finngen.gitbook.io/documentation

## Acknowledgements

We want to acknowledge the participants and investigators of the FinnGen study

